# Integration of family planning services into antiretroviral therapy for HIV in differentiated models of care in South Africa: a cross-sectional survey

**DOI:** 10.64898/2026.02.05.26345622

**Authors:** Oratile Mokgethi, Amy Huber, Idah Mokhele, Nyasha Mutanda, Vinolia Ntjikelane, Sydney Rosen, Musa Manganye, Lufuno Malala, Sophie Pascoe

## Abstract

**Introduction:** For differentiated models of care (DMOCs) that support client-centred HIV treatment (ART) in South Africa, a key next step in achieving integration is aligning clinic visits and medication dispensing for HIV treatment with other health needs like family planning. We assessed alignment between ART medication and family planning supply collection visits among DMOCs in South Africa.

**Methods:** We analysed self-reported data collected between September-December 2024 from women living with HIV (18-49 years, on ART ≥6 months) at 24 public healthcare facilities in four provinces (Gauteng, Mpumalanga, KwaZulu-Natal, and Eastern Cape). Participants were enrolled from four service delivery models: conventional care not eligible for DMOC (CN), conventional care eligible for DMOC but not enrolled (CE), facility pickup points (FAC-PuP), and external pickup points (EXT-PuP). Surveys assessed contraceptive use, visit alignment for injectable (Depo-Provera & Nur-Isterate) and oral contraceptive users, and how misaligned visits affected family planning adherence.

**Results:** Among 843 eligible women, 57% (460/843) reported current contraception use, with Depo-Provera being the most common (44%). Contraceptives users were younger (median 35 vs 38 years) and had slightly less ART experience (median 7 vs 8 years) than non-users. Contraceptive use varied by DMOC: CN (52%), CE (60%), FAC-PuP (63%), and EXT-PuP (50%). Half (131/260) of women using oral contraceptives or injectables collected their contraceptive and ART products on different days, with EXT-PuP showing the lowest level of alignment. Primary reasons for non-use were personal choice and beliefs (38%), followed by pregnancy-related factors (26%). Analysis of unmet family planning need in a subsample of 299 women found 22% had unmet need.

**Conclusion:** The findings reveal a high proportion of misalignment between ART and family planning services across models of care. Aligning ART and family-planning guidelines and services will promote ART and contraceptive adherence and reduce the burden on clients, maintaining the benefits of differentiated models and promoting integration of multi-condition service delivery.

## Introduction

Improving delivery of HIV care and of family planning represents a critical public health challenge globally, with particular significance in sub-Saharan Africa, where both HIV prevalence and unmet contraceptive needs remain high(1). Unmet need for family planning is defined as the proportion of women of reproductive age (15-49 years) who want to avoid or delay pregnancy but are not using contraception(2). South Africa, with an adult HIV prevalence of approximately 17%(3) and an unmet need for family planning of approximately 20% among all women of reproductive age (15-49 years)(4) –a figure that rises to over 50% among women living with HIV(5,6)—exemplifies this challenge(7). This context is particularly concerning given that an estimated 51.6% of pregnancies among women living with HIV are unplanned(5,8), making effective contraceptive access essential not only for eliminating mother-to-child HIV transmission but also for reducing adverse maternal and child health outcomes(7,9).

South Africa offers free contraceptive services and has the world’s largest HIV treatment program, currently reaching 75% of the HIV-positive population, with women comprising approximately two-thirds of those in treatment(3). Family planning services have been integrated into HIV treatment guidelines and procedures at health facilities for over a decade(10). Women seeking reproductive health services are counselled on HIV through provider-initiated counselling and testing (PICT), which is recommended as a standard component of medical care in all family planning settings(11). Conversely, guidelines call for women accessing HIV services to be offered contraceptive counselling and services as part of comprehensive care. Contraceptives are freely provided in all public health facilities regardless of the entry point. Despite these integration efforts, however, multiple barriers persist. These include structural challenges such as long waiting times and transportation difficulties(12,13), healthcare system factors including limited provider time for counselling and staff shortages(12), and individual-level barriers such as HIV-related stigma, cultural beliefs, partner preferences, and concerns about contraceptive side effects(12–14).

Like most other countries in the region (15,16), South Africa has adopted differentiated models of care (DMOCs) for HIV treatment that have significantly improved access to HIV care (17). Eligible HIV treatment clients who are enrolled in DMOCs can collect their antiretroviral therapy (ART) medications, other non-communicable disease medications, and some contraceptives such as oral contraceptives or short term injectables from facility-based (FAC-PuP) or external (community-based) pickup points (EXT-PuP), without attending full clinic visits (10,18). These models have successfully reduced barriers to chronic disease treatment access. They bring medication access closer to clients’ homes and workplaces, decongest facilities and reduce waiting times, and free up providers’ time for clients who need additional care (19,20).

Differentiated models of care present a significant opportunity for family planning integration, as aligning contraceptive dispensing with ART collection schedules would maximize the benefits of decentralized care while addressing persistent barriers to contraceptive access. The success of such integration, however, depends on compatible dispensing intervals between services, ensuring that contraceptive supplies can be provided on schedules that match ART collection and dispensing. We assessed the extent to which family planning and ART dispensing schedules are currently aligned and estimated numbers of missed doses and unmet family planning needs among adults on HIV treatment in South Africa, stratified by differentiated model of care.

## Methods

### Study setting and participants

This analysis uses data from the third round of SENTINEL, a multi-component observational study evaluating DMOC implementation in southern Africa (21). We report survey data collected between September 2024 and July 2025 at 24 public healthcare facilities in four South African districts (Ehlanzeni, Mpumalanga; Alfred Nzo, Eastern Cape; King Cetshwayo, KwaZulu Natal and West Rand, Gauteng). Study facilities were purposively selected based on ART client volumes, urban/rural distribution, and varying DMOC availability. SENTINEL has previously been described in detail(21).

Study participants were HIV positive adults (≥18 years) who had been receiving ART for at least six months and had experience with medication pickup in their respective model of care. For this analysis, we limited the analytic study sample to women aged 18-49 (reproductive age). Women who reported themselves to be sterile or post-menopausal were excluded from the study.

### HIV treatment delivery

In South Africa, ART delivery for virally suppressed clients occurs through three main differentiated models of care (DMOC): facility-based medication pickup points (FAC-PuP), external or community-based pickup points (EXT-PuP), and less commonly, adherence clubs. Eligibility for these models at the time of the study required at least 6 months on ART, documented viral suppression, and no active tuberculosis, pregnancy, or uncontrolled chronic conditions. Clients not enrolled in DMOCs, which included those eligible but not enrolled (CE) and those not eligible (CN), remained in conventional care(17). In this study, we define “conventional-eligible” and “conventional-not eligible” as two separate models of care.

Details of dispensing and visits for each model are presented in Supplementary Table 1. Dispensing durations for ART varied by model. Clients in conventional care received 2-3 month supplies of antiretroviral medications at each visit and were required to return for a full clinic visit each time they collected their medication, resulting in 4-6 full clinic visits per year. Once enrolled in a DMOC, clients continued to receive medication supplies of 2-3 months but only attended two full clinic visits per year at the facility. Between these clinic visits, they collected their ART refills at their selected pickup points. All clients were scheduled for viral load testing at 6 and 12 months after ART initiation, followed by annual testing as per South African treatment guidelines(22).

### Contraceptive service delivery according to South African national guidelines

In South Africa, women access modern contraceptive methods free-of-charge through the public healthcare system, each method a with distinct administration schedule and access pathway(10). Oral contraceptives (pills) require daily dosing and are typically dispensed in monthly supplies at healthcare facilities. However, women may also receive multi-month supplies (typically 2-3 months) to take home, reducing the need for monthly facility visits. Injectable contraceptives include two main options: Depo-Provera, a three-monthly injection, and Nur-Isterate (NET-EN), a two-monthly injection. Both are administered by nurses at primary healthcare facilities.

Male and female condoms are widely accessible through multiple distribution channels without requiring healthcare facility visits. Male condoms are available through community-based outlets (non-governmental organisations (NGOs), faith-based organizations), targeted venues (workplaces, public toilets, entertainment venues, truck stops, sports stadiums), and retail outlets (garages, pharmacies). Female condoms are distributed through community-based outlets and ward-based primary healthcare teams by trained community health workers, health promoters, and peer educators who can provide guidance on correct use.

Long-acting reversible contraceptive methods include subdermal implants (3-year protection), and intrauterine devices (IUDs) including both copper IUDs and the intrauterine system (IUS), which offer contraceptive protection for 5-10 years depending on the type. These methods require insertion by trained healthcare providers at primary healthcare clinics, after pregnancy is ruled out through clinical assessment.

### Study recruitment and consent

Data collectors received training on questionnaire administration prior to data collection. At each facility, we aimed to recruit up to 10 ART clients in each of the four models of care present at all study sites: CN, CE, FAC-PuP, and EXT-PuP. Participants were recruited sequentially upon arrival at the clinic for routine HIV-related care. Potential participants were screened for eligibility following referral from clinic staff and enrolled after providing written informed consent.

After consent, we administered a structured questionnaire that investigated, among other topics, current contraceptive use, alignment of ART and family planning visits, and how misaligned visits affected family planning adherence. These family planning survey questions were developed through literature review and consultations with expert family planning researchers and the SENTINEL research team. Relevant questions included several that were open-ended (qualitative).

During the data collection period, we recognized the importance of validating whether non-use of contraceptives reflected actual unmet need versus lack of demand. We thus expanded our survey instrument partway through data collection to include standardized unmet family planning need questions. These additional questions were administered to all eligible participants enrolled from that point forward, resulting in a consecutive subsample of 299 women who completed the extended questionnaire.

### Outcomes and analysis

The primary outcomes we report are: (1) prevalence of current self-reported contraceptive use stratified by DMOCs; (2) proportion of current non-users never offered contraceptive services; (3) alignment between contraceptive collection and ART visit schedules; and (4) unmet family planning need for a subsample of non-users. Table 1 presents the survey questions used to estimate each of these outcomes.

**Table 1.** Outcomes and relevant survey questions.

| Outcome | Survey questions related to outcome |
| --- | --- |
| Prevalence of current contraceptive use stratified by DMOCs | "Are you currently on any family planning method?" with response options including oral contraceptives, Depo injection, Nur-Isterate injection, condoms, implant, IUD/IUS, sterilization, emergency contraception, and other methods. Participants who selected 'Other' were asked to specify which method they were using.<br>Follow-up question: "If you are currently using contraceptives (oral, Depo or injectable) how frequently do you get a dose, refill or replacement of your contraceptive methods?" |
| Proportion of current non-users never offered contraceptive services | "If no, what are your reasons for not being on family planning?" and primary reasons for non-use were captured through the following structured response |
|  | categories:<br>○ **Personal choice and beliefs** (Religious/cultural reasons; I don't want to use contraception; Partner does not want me to use contraception)<br>○ **Pregnancy-related** (Currently pregnant; Planning pregnancy)<br>○ **Health and access concerns** (Fear of side effects; Because I can't get it at the same consultation as my ART; Never offered)<br>○ **Not currently sexually active** (derived from "Other" responses where clients specified they did not have a partner or were not sexually active).<br>▪ *"If you are not currently on contraceptives, are you offered contraceptives at your clinic visits when you see the nurse for your HIV review?"* |
| Alignment between contraceptive collection and ART visit schedules by DMOCs | ▪ *"When do you collect your contraception?"* with options: (1) "I always collect my ART medications and family planning on the same day," (2) "Sometimes it is on the same day as when I collect my ART medication but sometimes it is not," (3) "I always collect ART and family planning on different days."<br>○ Analysed and presented for women using oral contraceptives or injectable methods only |
| Proportion of missed doses asked to clients who reported always collecting on different days | ▪ *"Are you ever late or do you ever miss a dose of family planning because the schedule is different for ART collection and family planning?"* |
| Unmet family planning need (defined as sexually active, desired to delay or avoid pregnancy, but not using modern contraceptive methods) | ▪ Standardized unmet family planning need in a subsample of women using two key indicators. 1. Recent sexual activity "Have you been sexually active in the past 30 days?" 2. Fertility intentions "Thinking about your future plans, in the next 24 months, would you: (1) Want to have a child, (2) Want to delay or postpone having a child, (3) Do not want to have any (more) children, (4) Unsure/ Undecided about your plans, (5) Not applicable - Not of child bearing age." Unmet need defined as sexually active women who wanted to delay or avoid pregnancy but were not using modern contraceptive methods. |

**Table 2:**
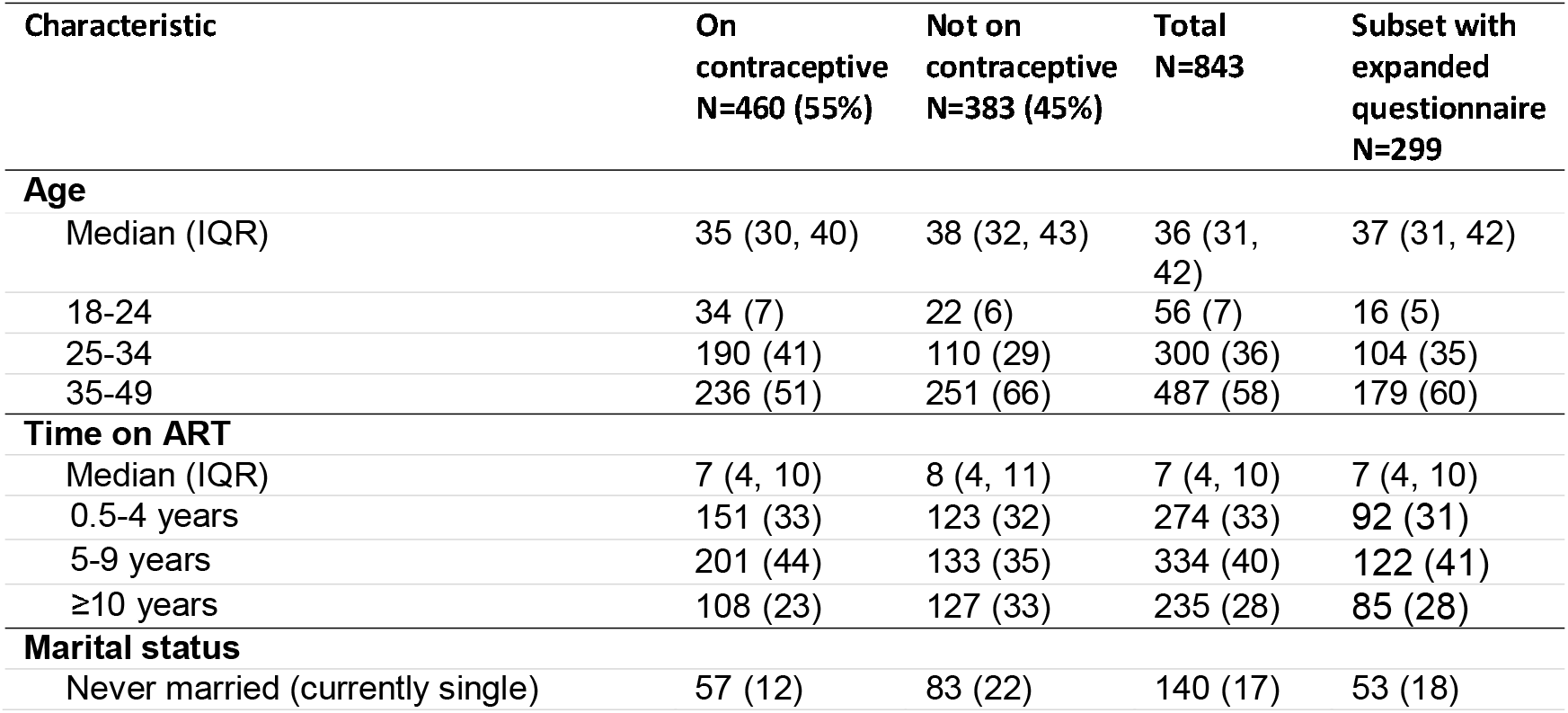
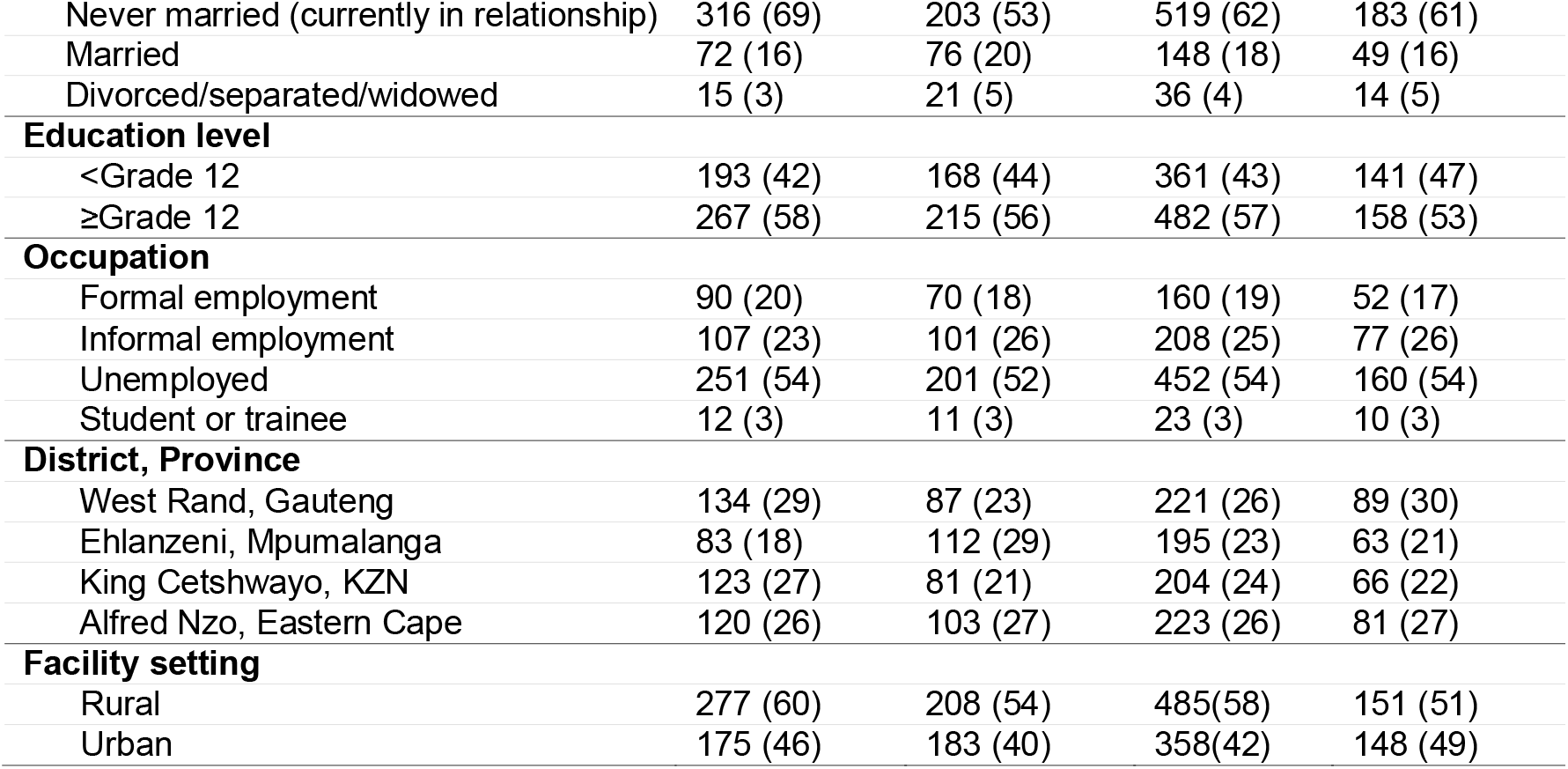
Demographic characteristics of women living with HIV by self-reported contraceptive use status in South Africa, 2024-2025.

| Characteristic | On<br>contraceptive<br>N=460 (55%) | Not on<br>contraceptive<br>N=383 (45%) | Total<br>N=843 | Subset with<br>expanded<br>questionnaire<br>N=299 |
| --- | --- | --- | --- | --- |
| **Age** |  |  |  |  |
| Median (IQR) | 35 (30, 40) | 38 (32, 43) | 36 (31, 42) | 37 (31, 42) |
| 18-24 | 34 (7) | 22 (6) | 56 (7) | 16 (5) |
| 25-34 | 190 (41) | 110 (29) | 300 (36) | 104 (35) |
| 35-49 | 236 (51) | 251 (66) | 487 (58) | 179 (60) |
| **Time on ART** |  |  |  |  |
| Median (IQR) | 7 (4, 10) | 8 (4, 11) | 7 (4, 10) | 7 (4, 10) |
| 0.5-4 years | 151 (33) | 123 (32) | 274 (33) | 92 (31) |
| 5-9 years | 201 (44) | 133 (35) | 334 (40) | 122 (41) |
| ≥10 years | 108 (23) | 127 (33) | 235 (28) | 85 (28) |
| **Marital status** |  |  |  |  |
| Never married (currently single) | 57 (12) | 83 (22) | 140 (17) | 53 (18) |
| Never married (currently in relationship) | 316 (69) | 203 (53) | 519 (62) | 183 (61) |
| Married | 72 (16) | 76 (20) | 148 (18) | 49 (16) |
| Divorced/separated/widowed | 15 (3) | 21 (5) | 36 (4) | 14 (5) |
| **Education level** |  |  |  |  |
| <Grade 12 | 193 (42) | 168 (44) | 361 (43) | 141 (47) |
| ≥Grade 12 | 267 (58) | 215 (56) | 482 (57) | 158 (53) |
| **Occupation** |  |  |  |  |
| Formal employment | 90 (20) | 70 (18) | 160 (19) | 52 (17) |
| Informal employment | 107 (23) | 101 (26) | 208 (25) | 77 (26) |
| Unemployed | 251 (54) | 201 (52) | 452 (54) | 160 (54) |
| Student or trainee | 12 (3) | 11 (3) | 23 (3) | 10 (3) |
| **District, Province** |  |  |  |  |
| West Rand, Gauteng | 134 (29) | 87 (23) | 221 (26) | 89 (30) |
| Ehlanzeni, Mpumalanga | 83 (18) | 112 (29) | 195 (23) | 63 (21) |
| King Cetshwayo, KZN | 123 (27) | 81 (21) | 204 (24) | 66 (22) |
| Alfred Nzo, Eastern Cape | 120 (26) | 103 (27) | 223 (26) | 81 (27) |
| **Facility setting** |  |  |  |  |
| Rural | 277 (60) | 208 (54) | 485(58) | 151 (51) |
| Urban | 175 (46) | 183 (40) | 358(42) | 148 (49) |

We used descriptive statistics and frequency distributions to examine sociodemographic and clinical factors, stratified by contraceptive use status. Contraceptive use patterns, including method type and service integration alignment, were analysed by DMOCs. For the unmet need subsample, we calculated prevalence of unmet family planning need overall and by DMOC, then cross-referenced these findings with contraceptive offering patterns to validate service gap identification.

### Ethics review

The SENTINEL study was approved by the Human Research Ethics Committee (Medical) of the University of Witwatersrand (South Africa), protocol number M210241 and the Boston University IRB (United States), protocol numbers H-41402. All participants provided written informed consent for the study procedures.

## Results

### Sample selection

Study enrolment is illustrated in Figure 1. Between September 2024 and July 2025, we enrolled 1,460 participants in SENTINEL 3.0 and retained 843 for this analysis. The parent study enrolled both male and female clients; males (n=360) were excluded from this sub-study focused on family planning needs.

**Figure 1:**
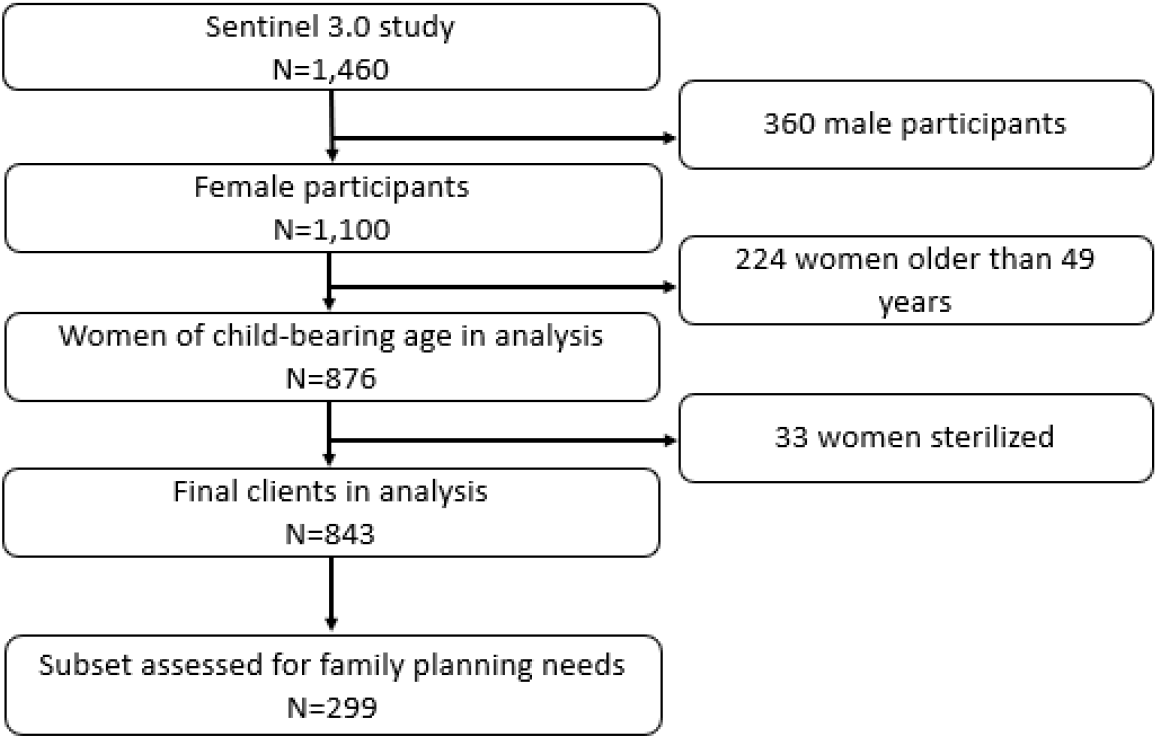
Study enrolment. Demographic characteristics

Of the 843 women in this analysis, 460 (55%) reported that they were currently using some form of contraception at the time of the survey. Women using contraceptives were slightly younger than those who were not (median age 35 years vs 38 years), with more in the 25-34 age group (41% vs 29%). Time on ART likely reflected this age difference, with non-users more likely to have been on treatment for ten or more years (33% v 23%). Contraceptive users were more likely to be in relationships but never married (69% vs 53%), while non-users were more likely to be single (22% vs 12%). Contraceptive use differed by location, with users representing 29% of participants from West Rand, 27% from King Cetshwayo, 26% from Alfred Nzo, and 18% from Ehlanzeni. Contraceptive users showed marginally lower representation in the conventional care ineligible (CN) model than did non-users (49% vs 53%). There were small differences in differentiated model enrolment: users were marginally more likely to use facility pick-up points (15% vs 11%) while non-users showed slightly higher use of external pick-up points (21% vs 17%).

### Contraceptive use patterns by differentiated service delivery model

The likelihood of contraceptive use varied by differentiated model of care, as mentioned above. Table 3 reports methods of contraception used and reasons for non-use, by model of care. Injectable contraceptives were the predominant method across all service delivery models, accounting for 51% of contraceptive use overall (44% Depo injection and 7% Nur-Isterate). Condoms (21%) and implants (19%) were both used by about a fifth of the sample, while oral contraceptives demonstrated consistently low uptake across all groups (5%). There were modest differences in use of different methods by model of care, with clients enrolled in external pickup points slightly more likely to rely on condoms and less likely to use Depo injections than in other models.

**Table 3:** Self-reported contraceptive use, contraception offer, and reasons for non-use among women living with HIV, by model of care.

| Characteristic | Conventional,<br>not eligible<br>(CN) | Conventional,<br>eligible but not<br>enrolled (CE) | Facility<br>pick-up<br>point<br>(FacPuP) | External<br>pick-up<br>point<br>(ExPuP) | Total |
| --- | --- | --- | --- | --- | --- |
| N (n, row %) | 427 (51) | 147 (17) | 112 (13) | 157 (19) | 843 (100) |
| On contraceptives (n, % of model) | 223 (52) | 88 (60) | 71 (63) | 78 (50) | 460 (55) |
| Method of contraception (n, column %) |  |  |  |  |  |
| Oral contraceptives (pills) | 9 (4) | 5 (6) | 4 (6) | 6 (8) | 24 (5) |
| Injection- Depo (3-month) | 99 (45) | 46 (52) | 30 (42) | 28 (36) | 203 (44) |
| Injection – Nuristerate (2-month) | 11 (5) | 5 (6) | 9 (13) | 8 (10) | 33 (7) |
| Condoms | 48 (21) | 17 (19) | 14 (20) | 20 (26) | 99 (21) |
| Implant | 52 (23) | 12 (14) | 13 (18) | 11 (14) | 88 (19) |
| IUD and IUS (loop) | 4 (2) | 3 (3) | 1 (1) | 4 (5) | 12 (3) |
| Other | 0 | 0 | 0 | 1 (1) | 1 (0) |
| Not on contraceptives (n, % of model) | 204 (48) | 59 (40) | 41 (37) | 70 (50) | 383 (45) |
| Offered contraceptive if not currently using (n, % of model not on contraceptives) | 51 (25) | 19 (32) | 8 (20) | 21 (30) | 99 (26) |
| Reasons for non-use (n, column %) |  |  |  |  |  |
| Personal choice and beliefs | 65 (32) | 21 (36) | 16 (39) | 44 (56) | 146 (38) |
| Pregnancy-related | 77 (38) | 5 (8) | 9 (22) | 7 (9) | 98 (26) |
| Health and access concerns | 45 (22) | 26 (44) | 10 (24) | 20 (26) | 101 (26) |
| Not currently sexually active | 13 (6) | 5 (8) | 4 (10) | 6 (8) | 28 (7) |
| Other | 4 (2) | 2 (3) | 2 (5) | 1 (1) | 9 (2) |

Among the 383 (45%) women not using contraceptives, only 99 (26%) reported being offered contraceptive services, while the majority (74%) said they had not been offered contraceptives. The proportion of women offered contraceptives varied by service delivery model, with those eligible for but not enrolled (CE) reporting the highest rate of contraceptive offer (32%), compared to 30% among external pick-up point users, 25% among conventional care not eligible recipients (CN), and 20% among facility pick-up point users.

The most common reasons for contraceptive non-use were personal choice and beliefs (38%), followed by pregnancy-related factors (26%) and health and access concerns (26%). Reasons for non-use varied considerably by model of care. Women using external pick-up points were most likely to cite personal choice and beliefs as their primary reason for non-use (56%), while those in conventional care were more likely to report pregnancy-related reasons (38%). Health and access concerns were most prevalent among women eligible for but not enrolled in DMOC (44%), suggesting perceived or real barriers to contraceptive access within this group. Women who self-reported not being sexually active represented a small proportion of the sample (7% overall), with relatively consistent rates across service delivery models.

### Self-reported alignment of HIV treatment and contraceptive collection by DMOC

Half of the participants using oral contraceptives and injectables (131/260, 50%) said that their medication collection schedules were not aligned: they collected their contraceptive supplies and ART medications on different days (Figure 2). Alignment varied by model of care. Women not eligible for DMOC and those eligible but not enrolled had the highest same-day collection rates at 29% and 32% respectively. Those enrolled in facility pick-up points had lower alignment, with approximately 23% collecting ART and family planning services on the same day and an additional 40% having mixed scheduling patterns (sometimes collecting on the same day, sometimes on different days). External pick-up point users had the lowest same-day collection rate at approximately 7%, with the majority (approximately 60%) collecting ART and family planning services on different days, likely requiring engagement with different service locations.

**Figure 2:**
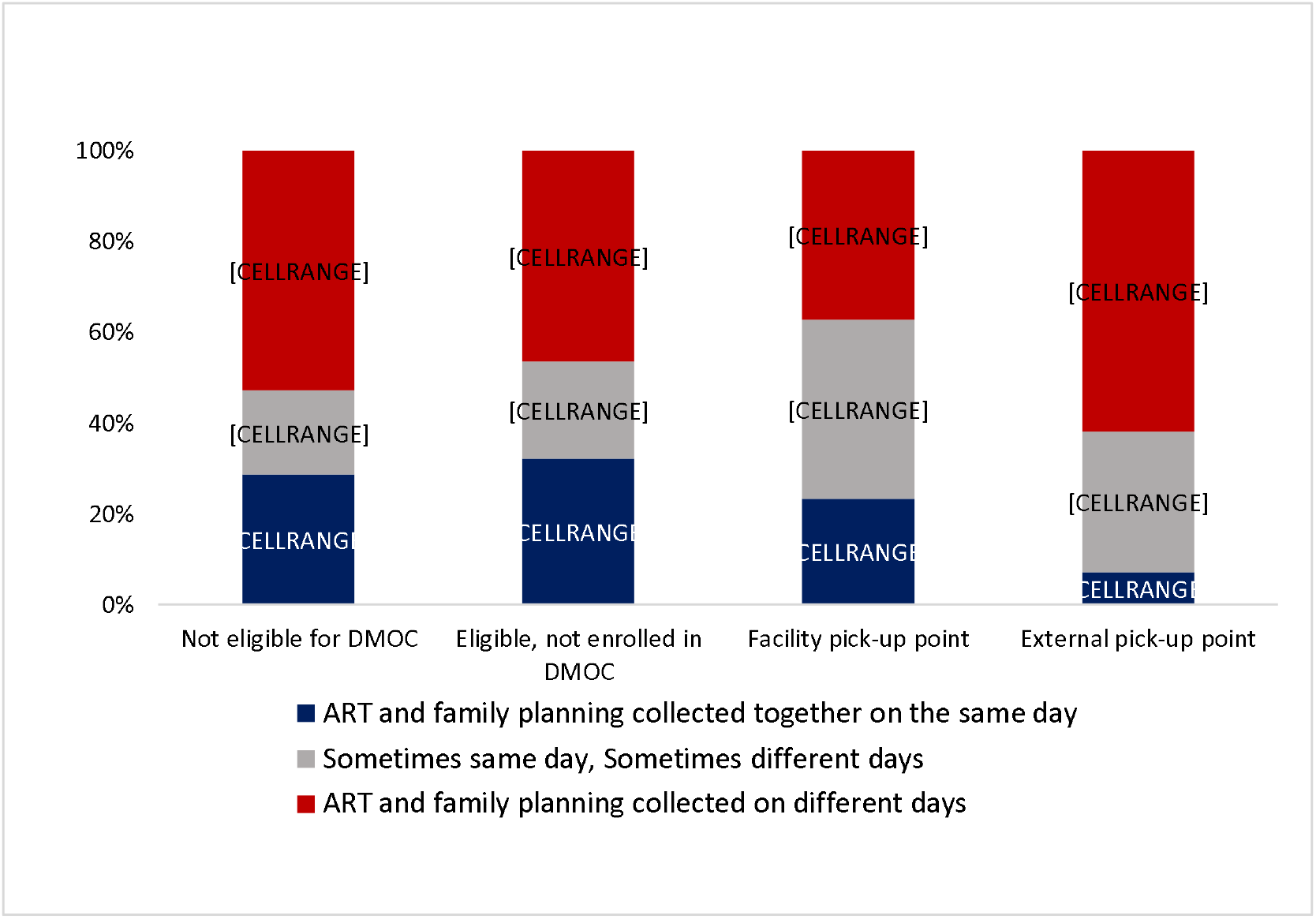
Self-reported alignment of contraceptive collection schedules with ART visits among women using oral contraceptives and injectables, by model of care.

### Missed doses resulting from misalignment of visits

Among the 260 women using oral contraceptives or injectable methods, 24 (9%) reported a late or missed dose of family planning that they attributed to misaligned medication pickup schedules. Three main themes emerged explaining how misaligned schedules affected contraceptive adherence (Table 4). Women described managing separate appointments that led to forgetfulness, with some noting confusion from having multiple appointment cards or completely forgetting family planning visit dates. Financial barriers also prevented attendance at separate appointments, as women cited concerns about losing daily wages, transport costs, and the economic burden of making multiple trips to healthcare facilities. Some participants observed that collecting both medications on the same day was more cost-effective, and some reported strategically delaying contraceptive supply pickup to align appointments, particularly when appointment dates were close together, with some intentionally timing their visits to receive both services simultaneously or waiting until they had sufficient contraceptive supplies to cover the gap until their next ART visit.

**Table 4:**
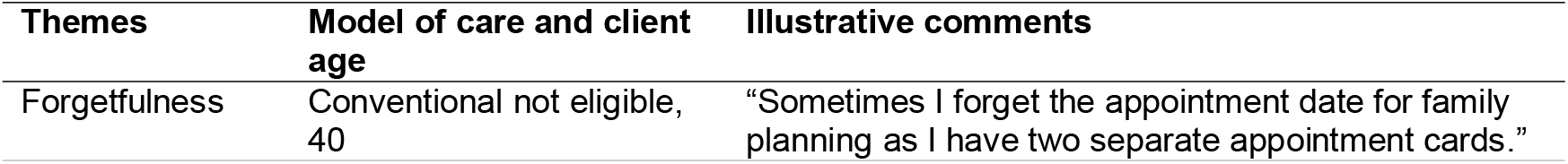
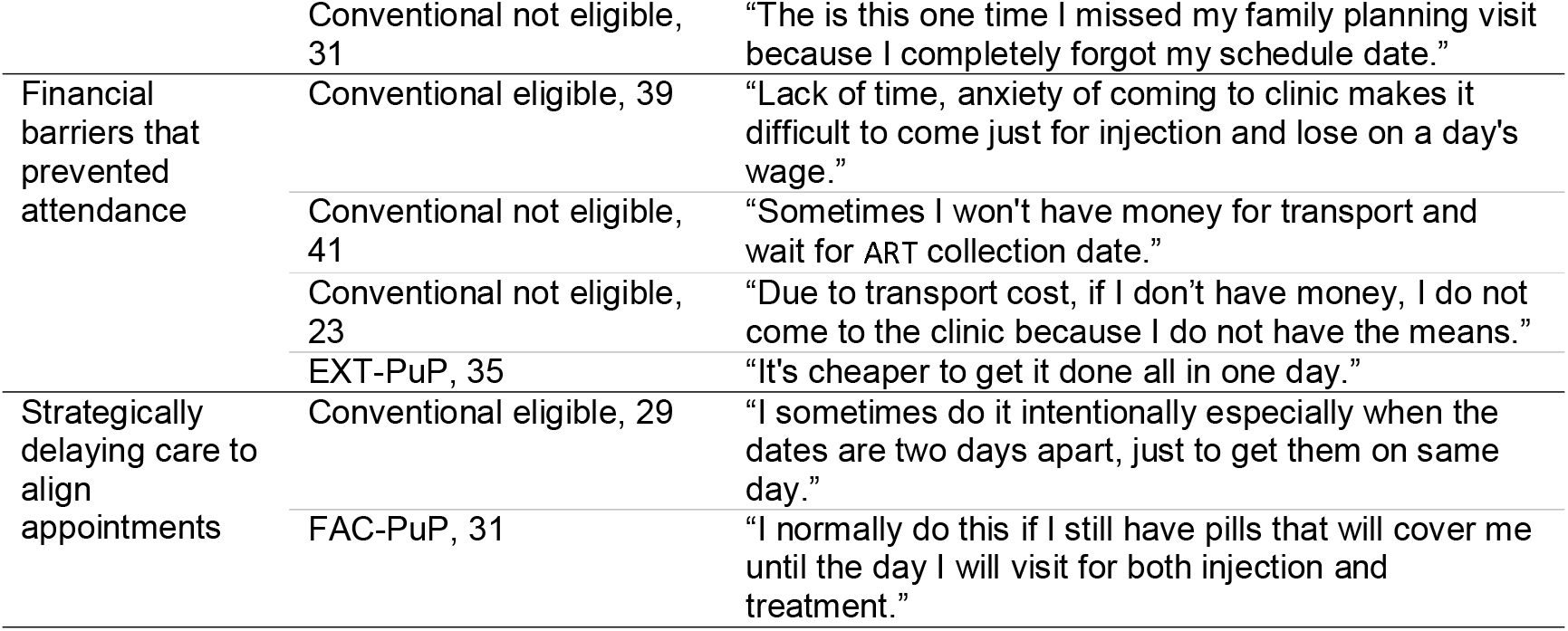
Participant experiences of misaligned contraceptive and ART collection schedules.

**Table 4:** Unmet family planning need, by model of care (n=299)

| Model of care (n, row %) | No unmet need | Unmet need | Total |
| --- | --- | --- | --- |
| N | 263 (88) | 36 (12) | 299 (100) |
| Conventional, not eligible for DMOC (CN) | 139 (89) | 17 (11) | 156 (100) |
| Conventional, eligible for DMOC (CE) | 33 (80) | 8 (20) | 41(100) |
| FAC-PuP | 42 (95) | 26 (5) | 44 (100) |
| EXT-PuP | 49 (84) | 9 (16) | 58 (100) |

### Unmet family planning needs assessment

To explore whether contraceptive non-use was associated with unmet need, we assessed family planning need in a subsample of 299 women. Among these, 148 /299(49%) were not using contraceptives and completed both sexual activity and fertility intention questions. Of the 148 non-contraceptive users, 87 (59%) reported being sexually active in the past 30 days. Among the 87 sexually active non-users, 36 (41%) expressed a desire to delay or avoid pregnancy. Unmet need for family planning - defined as not using contraception while being sexually active and wanting to delay or avoid pregnancy - thus affected 36 women, representing 12% of the total subsample (36/299).

Unmet need varied significantly by model of care (Table 5), with the highest proportion (20%) among women eligible to be enrolled in DMOCs, compared to 5% among those using facility pickup points. There was no difference in the likelihood of unmet need between those who said that they were or were not offered contraception by the clinic. The majority of non-users (72%) in both groups had never been offered contraceptive services.

## Discussion

In this study of contraceptive use patterns and the alignment of family planning and ART collection schedules among women living with HIV and on treatment in four districts in South Africa, we found that 55% of reproductive-age women were currently using modern contraception and that exactly half of contraceptive users reported that their medication collection schedules were not aligned: they collected their contraceptive supplies and ART medications on different days. This misalignment had practical consequences, with some women reporting missed contraceptive doses due to separate appointment systems, financial barriers due to multiple facility visits, and logistical challenges in managing different collection schedules. Among those not on contraceptives, only a quarter (26%) reported ever having been offered contraception by their clinic.

The predominance of injectable contraceptives observed in our study, particularly three-monthly Depo-Provera (44% of users), is consistent with findings from other sub-Saharan African countries, where injectable methods are often preferred due to their discretion, effectiveness, reduced need for daily pill adherence, and partner involvement(23,24). The relatively high uptake of implants (19% of users) suggests growing acceptance of long-acting reversible contraceptive methods(25), which could be ideal for women in DMOCs due to their minimal follow-up requirements. Conversely, the low uptake of oral contraceptives (5% of users) likely reflects multiple challenges including the burden of daily adherence and monthly refill requirements that conflict with extended ART dispensing intervals(26).

The high proportion of women not currently using contraception who reported never being offered contraceptive services (74%) highlights a gap in integrated counselling and service delivery, despite South Africa’s longstanding policies promoting HIV care-family planning integration(10,27). This finding reflects broader implementation challenges documented in the literature. Despite supportive integration policies, actual service delivery often remains vertical, with referrals between services rather than true integration(28). As noted by Kriel et al., discussions surrounding family planning integration with primary healthcare services in South Africa revealed several operational challenges. Clinicians providing HIV services are not always trained in family planning provision, nor do they routinely have contraceptives available at the point of HIV care, making it difficult to organize efficient services that accommodate diverse client needs(28). Participants in their study also reported that integration actually negatively affected contraceptive access in some settings, while another study found that despite service integration efforts, unplanned pregnancy rates remained very high(29).

The disconnect between policy intent and ground-level implementation appears to extend to our study context as well, where contraceptive counselling is not systematically offered across differentiated HIV care models. Our analysis of unmet need reveals that the challenge extends beyond simply offering services. Even when contraceptive counselling is provided, significant barriers to uptake remain. The persistence of unmet need, even among counselled women, suggests that there are underlying structural, financial, and individual-level barriers that prevent contraceptive uptake and continuation for this HIV-positive population.

Contraceptive use varied across different care models, suggesting that the HIV care approaches may influence family planning access and utilisation. In facility-based models where infrastructure for service integration exists (10)—i.e., in FAC-Pup and both models of conventional care--we expected to see a high proportion of same-day ART and contraceptive collection. The majority of participants in these models reported collecting sometimes or always on different days, however, indicating lack of coordination between ART and family planning services within clinics (28).

The challenges are more pronounced for clients in external pickup points (Ex-PuP), where nearly two-thirds of women using injectables or oral contraceptives collected them on completely different days from their ART. This separation is partly structural, as injectable contraceptives in South Africa are provided only within health facilities by trained nurses, creating an inherent disconnect from community-based ART collection points. The 3-month administration intervals for injectable contraceptives require quarterly clinic visits regardless of ART dispensing intervals, reducing the client benefits of community ART pickup points (EXT-PuP). From the client’s perspective, there is little value in collecting HIV medications from an external pickup point if dispensing could be aligned at the facility. We note that condom use was slightly higher among women in external pickup points than in other models of care, which may reflect the convenience of using contraception that can be obtained at these community-based sites alongside ART collection(26,30).

When ART and family planning visits occur on separate schedules, women are forced to make multiple facility visits, incur additional transport costs, and manage complex appointment systems-- effectively recreating the very barriers that differentiated service delivery models for HIV were meant to eliminate. Some women reported strategically delaying care to align appointments, and many experienced financial hardships from repeated travel costs. These financial barriers are observed in similar contexts, where male participants have noted that the financial burden of accessing contraception often falls on them due to women’s economic dependence, as seen in studies examining couple perspectives on contraceptive access(28). These barriers ultimately affect individual autonomy by constraining women’s independence to make autonomous reproductive choices that should be based on personal preferences and health needs rather than financial constraints.

This study has several limitations that should be considered when interpreting the findings. The cross-sectional design limits our ability to assess causality or changes over time. Self-reported data may be subject to social desirability bias, particularly regarding contraceptive use and service experiences. The study was conducted at selected facilities and may not be representative of all DMOC implementation sites in South Africa. Additionally, we did not assess provider perspectives or health system factors that may influence service delivery, limiting our understanding of implementation barriers. The unmet family planning need assessment was conducted in only a subset of participants (n=299) due to mid-study questionnaire modifications, which may limit the generalizability of these specific findings. Furthermore, our definition of unmet need, while based on standard demographic health survey criteria, may not capture the full complexity of contraceptive decision-making.

## Conclusion

For policymakers, our findings demonstrate that South Africa’s successful DMOC program represents an untapped opportunity for comprehensive reproductive health integration. These results, when considered alongside the country’s goals to eliminate mother-to-child HIV transmission and reduce unplanned pregnancies, suggest that policymakers and program managers should prioritize re-training HIV care providers in integrated contraceptive counselling and point-of-care administration of compatible methods, introducing self-administered contraceptive options that better align with decentralized care schedules, and expanding contraceptive service delivery to community-based pickup points.

## Data Availability

All data produced in the present study are available upon reasonable request to the authors

## Supplementary files

Supplementary table 1. Differentiated service delivery models and conventional care characteristics

## Authors’ contributions

Study conceptualization was led by NOM, AH, IM, NM, SP, and SR. The research instrument was collaboratively developed by AH, IM, NM, SP and SR. Data collection was supervised by AH and VN, while NM oversaw the data cleaning process. NOM conducted the data analysis and prepared the initial manuscript draft, incorporating inputs from AH, IM, NM, VN, SP, LM, MM, and SR. All authors participated in the manuscript review and revision process prior to submission.

## Data availability statement

All data used in this study were collected by the study team following written informed consent. Data will be made publicly available in a data repository within one year of study closure, as approved by the supervising ethics committees.

