## Supplementary Table 1 for "Integration of family planning services into antiretroviral therapy for HIV in differentiated models of care in South Africa: a cross-sectional survey"

**Supplementary table 1. Differentiated service delivery models and conventional care characteristics**

| **Model** | **Model Description** | **Medication supply** | **Annual facility visits** | **Other interactions** |
| --- | --- | --- | --- | --- |
| Conventional care – eligible for DMOC | Clients eligible for differentiated models but remained in conventional care due to declining enrollment or facility/provider limitations | 2-3-month supply of medications  at each full clinic visit | 4-6 facility visits | 0 |
| Conventional care – not eligible for DMOC | Clients who remained in conventional care because they did not meet DMOC eligibility criteria | 2-3-month supply of medications  at each full clinic visit | 4-6 facility visits | 0 |
| Facility based pickup-points (FAC-PuP) | Pre-packaged medications collected at facility without full clinic visit | 2-3-month supply of medications  at each full clinic visit | 2 facility visits | 2-4 medication  pick ups |
| External pickup-points (EXT- PuP) | Medications collected at community pickup points (e.g., commercial pharmacy, adherence clubs) * | 2-3-month supply of medications  at each full clinic visit | 2 facility visits | 2- 4 medication  pick ups |

*Adherence clubs in the Eastern Cape were grouped under EXT-PuP for analysis due to small numbers
